# Biallelic truncation variants in *ATP9A* are associated with a novel autosomal recessive neurodevelopmental disorder

**DOI:** 10.1101/2021.05.31.21257832

**Authors:** Francesca Mattioli, Hossein Darvish, Sohail Aziz Paracha, Abbas Tafakhori, Saghar Ghasemi Firouzabadi, Marjan Chapi, Hafiz Muhammad Azhar Baig, Alexandre Reymond, Stylianos E. Antonarakis, Muhammad Ansar

**Affiliations:** Center for Integrative Genomics, University of Lausanne, Lausanne, Switzerland; Neuroscience Research Center, Faculty of Medicine, Golestan University of Medical Sciences, Gorgan, Iran; Anatomy Department, Khyber Medical University Institute of Medical Sciences (KIMS) Kohat, Pakistan; Iranian Center of Neurological Research, Neuroscience Institute, Tehran University of Medical Sciences, Tehran, Iran; Genetics Research Center, University of Social Welfare and Rehabilitation Sciences, Tehran, Iran; Department of Biochemistry, Institute of Biochemistry, Biotechnology and Bioinformatics, The Islamia University of Bahawalpur, Bahawalpur, Pakistan; Department of Genetic Medicine and Development, University of Geneva Medical Faculty, Geneva 1211, Switzerland; Medigenome, Swiss Institute of Genomic Medicine, Geneva, Switzerland; Institute of Molecular and Clinical Ophthalmology Basel (IOB), Basel, Switzerland

**Keywords:** *ATP9A*, Autosomal Recessive Intellectual Disability, Consanguinity, Runs of Homozygosity

## Abstract

Intellectual disability (ID) is a highly heterogeneous disorder with hundreds of associated genes. Despite progress in the identification of the genetic causes of ID following the introduction of high-throughput sequencing, about half of affected individuals still remain without a molecular diagnosis. Consanguineous families with affected individuals provide a unique opportunity to identify novel recessive causative genes.

In this report we describe a novel autosomal recessive neurodevelopmental disorder. We identified two consanguineous families with homozygous variants predicted to alter the splicing of *ATP9A* which encodes a transmembrane lipid flippase of the class II P4-ATPases. The three individuals homozygous for these putatively truncating variants presented with severe ID, motor and speech impairment, and behavioral anomalies. Consistent with a causative role of *ATP9A* in these patients, a previously described *Atp9a-/-*mouse model showed behavioral changes.

## INTRODUCTION

Intellectual disability (ID) or delayed psychomotor development are common and highly heterogeneous phenotypes of genetic origin, affecting 1-3% of the general population ^1,2^ which pose a significant socio-economic burden on the affected families, the health care system, and society in general ^3^. Despite considerable progress in genetic diagnosis after the introduction of high throughput sequencing technologies, the genetic cause of more than half of ID cases remains undetermined ^4^. The leading genetic cause of ID in individuals from outbred populations is *de novo* variants ^5,6^; in contrast a substantial fraction of autosomal recessive (AR) disorders cause ID in families with multiple affected individuals that practice consanguinity ^7^. It is estimated that worldwide 10.4% of marriages occur among close relatives ^8^. Consanguinity increases the extent of homozygous genomic regions and brings to homozygosity deleterious alleles resulting in birth defects and infant mortality ^9,10^. Large consanguineous families with (multiple) affected individuals thus provide a unique opportunity to identify novel recessive causative genes.

P4-ATPases are transmembrane lipid flippases ^11^, that function in vesicles formation and trafficking. They regulate the asymmetric distribution of phospholipids in membranes of eukaryotic cells ^11,12^. There are 14 different P4-ATPases in humans that can be phylogenetically grouped in five classes^13^. ATP9A and its 75% similar paralog ATP9B are the unique members of class II. They are the only P4-ATPase that do not require the CDC50 β-subunit for normal function and cellular localization ^14^. They show different intracellular and tissue distribution: ATP9A is found in early and recycling endosomes and at a lower level at the plasma membrane, while ATP9B is only found in the trans-Golgi network ^12,14–16^. Similarly, the genes encoding ATP9A and ATP9B present with overlapping but different expression patterns with *ATP9A* mainly expressed in the brain (Human Protein Atlas, GTEx). Suggestive of an important role of ATP9A in intercellular communication, this P4-ATPase inhibits extracellular vesicles release ^15,16^.

Here we report two consanguineous families with homozygous pathogenic variants predicted to alter *ATP9A* splicing and we propose ATP9A as a novel cause of a recessive neurodevelopmental disorder.

## RESULTS

### CLINICAL REPORT

We identified three affected individuals from two unrelated consanguineous families. The main clinical features of the affected individuals are reported in Table 1 and in Figure 1. Because rapid and automated nature of preprint posting are incompatible with verification of informed consent and that data in this section can potentially make patients identifiable, the clinical reports of the affected individuals are available upon request to the corresponding authors.

**Table 1:** Clinical features of patients with homozygous *ATP9A* splicing variants

| **Family** | **1** |  | **2** |
| --- | --- | --- | --- |
| **Individual** | **IV:1** | **IV:7** | **IV:2** |
| Consanguineous parents | yes | yes | Yes |
| Age at last evaluation (bracket of years) | 25-30 | 20-25 | 10-15 |
| *ATP9A* variant (gDNA) | Chr20:50305602 C>A | Chr20:50305602 C>A | Chr20:50342357 C>A |
| ***General characteristic*** |  |  |  |
| Head circumference (cm) | <1st percentile | 39<sup>th</sup> percentile | 50<sup>th</sup> percentile |
| Height (cm) | <3rd percentile | 50-75<sup>th</sup> percentile | 50-75<sup>th</sup> percentile |
| BMI (range) | 25-29 | 20-24 | 20-24 |
| Microcephaly | + | - | - |
| Strabismus | + | + | - |
| Facial dysmorphism | + | + | + |
| ***Neurodevelopment*** |  |  |  |
| Severe Intellectual Disability | + | + | + |
| Motor delay | + | + | + |
| Speech delay/ dysfunction | + | + | + |
| Fine motor impairment | + | + | + |
| Epilepsy | - | - | + |
| Brain MRI anomalies | n.d. | n.d. | - |
| ***Behavioral anomalies*** |  |  |  |
| ADHD | + | + | n.d. |
| Stereotypic movement | n.d. | n.d. | + |
| Autistic features | - | - | + |
| Aggressiveness | + | + | n.d. |

n.d. = not determined; ADHD = Attention Deficit Hyperactivity Disorder

**FIGURE 1:**
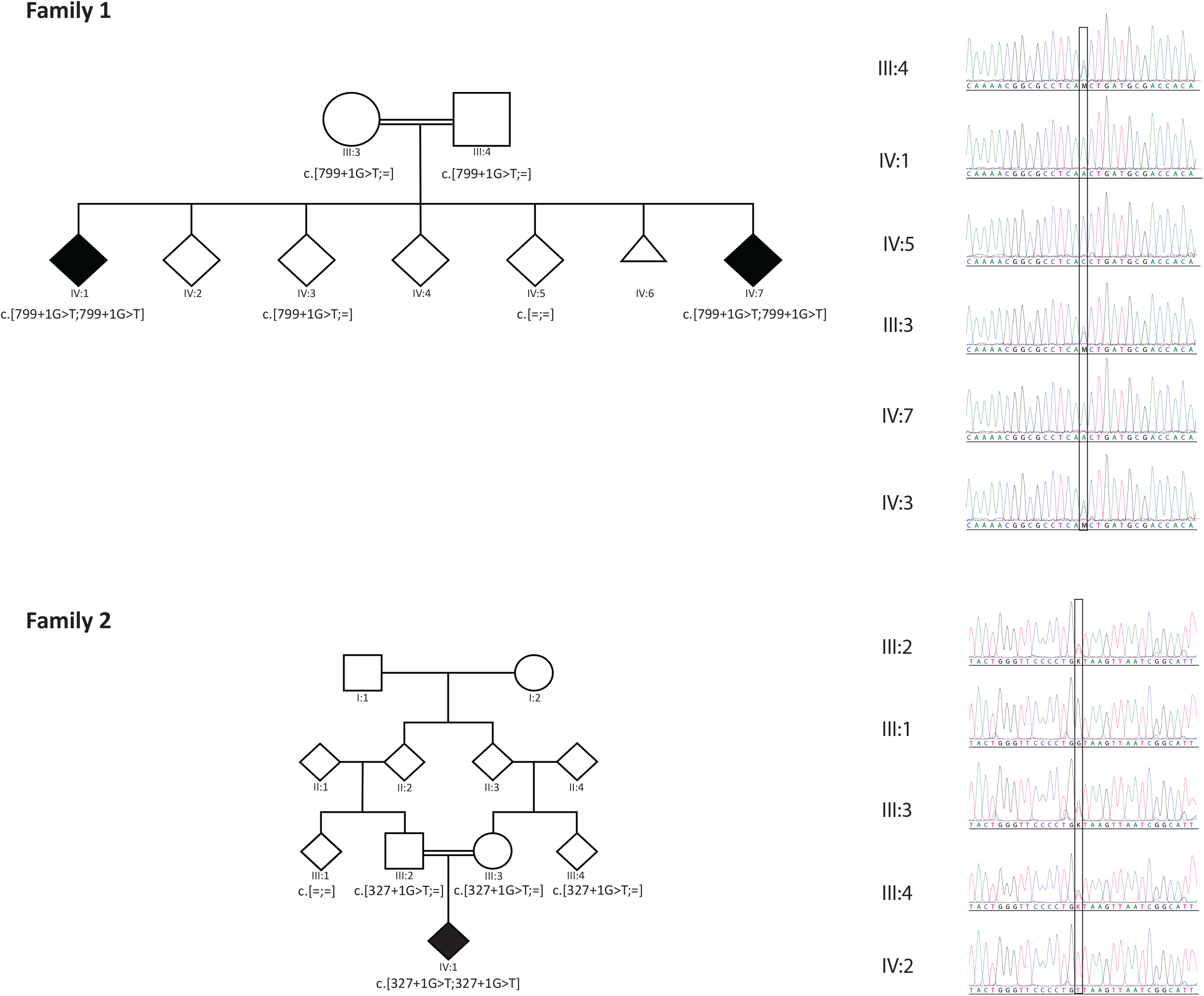
Pedigrees and Sanger sequencing. The pedigrees and the available genotypes of family 1 (top) and family 2 (bottom) are depicted on the left. Sanger sequencing chromatograms confirming the segregation of the *ATP9A* NM_006045.3:c.799+1G>T (six top traces) and NM_006045.3:c.327+1G>T variants (bottom five traces) are shown on the right.

### EXOME ANALYSIS

Homozygosity mapping of family 1 and whole exome sequencing (WES) of proband (IV:1) of family 1 did not reveal any pathogenic or likely pathogenic mutation in genes previously reported to cause ID or developmental delay but allowed the detection of a homozygous splicing variant (NM_006045.3:c.799+1G>T) in *ATP9A* (Figure 1). The variant was not present in gnomAD^17^, Bravo (https://bravo.sph.umich.edu/freeze5/hg38/) or our local database of >500 controls from the same geographical region. Its segregation in the family was confirmed by Sanger sequencing, in particular the two affected siblings are homozygous for this variant (Figure 1). The change at the conserved first nucleotide of the donor splice site was predicted to cause abnormal splicing by SpliceAI ^17^ (score DS_DL = 0.99), MaxEntScan^18^ (MaxEntScan_diff = 8.504), and NNsplice^19^. RNA samples from affected individuals were not available to assess RNA splicing.

Our search for more cases led to the identification of a second family. The WES of proband IV:1 from family 2 also revealed the presence of a homozygous splicing variant in *ATP9A*, a base pair substitution in intron 3 of *ATP9A* (NM_006045.3:c.327+1G>T;). This variant is absent from the gnomAD^17^ and Bravo databases, and from a database of hundreds of healthy individuals and our local database of >250 controls from the same geographical region. Multiple predictions tools indicated a likely loss of the canonical donor splice site (NNsplice, SpliceAI score DS_DL=0.95, MaxEntScan_diff = 8.504). The abnormal splicing could either result in the skipping of the inframe exon 3, leading to the deletion of 38 amino acid residues, or utilization of an alternative donor site resulting in partial intronic retention and the appearance of a premature stop codon. Testing of the aberrant RNA splicing was not possible due to unavailability of patient’s RNA or cells. Sanger sequencing confirmed the segregation of the potentially causative variant (Figure 1), i.e. the variant is heterozygous in the proband’s parents (III:2 and III:3). Homozygosity mapping of the proband revealed that the *ATP9A* variant is embedded in a putative 6.83 Mb region of homozygosity (ROH) (chr20[GRCh37]: 45358223-52192534). While we did not find any likely-pathogenic variants in known ID genes in proband IV:1 of family 2 (based on the Panelapp gene list for intellectual disability ^20^; Supplementary Table 1), we cannot exclude that variants beside the *ATP9A* one might play a role in the patient’s phenotype. In particular, we identified homozygous variants in *CCDC88C* (NM_001080414.4: c.1126C>T, p.Arg376Trp) and *ZNF407* (NM_017757.3: c.5497>T, p.Pro1833Ser), two genes previously implicated in neurodevelopmental disorders but associated with phenotypes different than the one found in our proband. Bi-allelic variants in *CCDC88C* were associated with a form of congenital hydrocephalus ^21–23^, while variant in *ZNF407* have been recently implicated in a autosomal recessive form of ID with microcephaly, short stature, hypotonia and ocular anomalies ^24,25^.

## DISCUSSION

Autosomal recessive ID is characterized by extensive genetic heterogeneity. Still, many patients do not receive a molecular diagnosis, suggesting that a considerable number of causative genes have not yet been identified ^4,27^. We described three individuals from two consanguineous families with different homozygous splicing variants in canonical splice sites of the *ATP9A* gene. All three patients present with severe ID, motor delay, speech and fine motor impairment and behavioral anomalies. Both affected siblings (IV:1 and IV:7) of family 1 had an attention deficit hyperactivity disorder-like phenotype combined with aggressiveness, whereas proband IV:1 from family 2 presented with autistic features, including prominent stereotypic movements, and lack of eye contact.

*ATP9A* is under constraint (intolerance to missense variants z-score = 4.15; pLI = 1; LOEUF = 0.2) according to gnomAD^26^. Its yeast homolog, *NEO1*, was shown to be an essential gene ^28^, while absence of the *C. elegans* orthologous *TAT-5* resulted in disrupted cell adhesion and morphogenesis in worms’ embryos ^29^. Whereas ablation of the mouse orthologous *Atp9a* did not diminish survival, the *Atp9a*^*-/-*^ mice engineered and phenotyped by the International Mouse Phenotyping Consortium were hyperactive and showed a significant increased exploration in new environment reminiscent of the behavioral symptoms of our patients ^30,31^. Depletion of *ATP9A* were lethal in human hepatoma HepG2 cells but not in other cell lines including HeLa, HEK293T, MCF-7, and THP-1, suggesting that the absence of ATP9A could be tolerated in certain tissues but not in others ^12,15^. *ATP8A2*, another P4-ATPase highly expressed in the brain, has been implicated in a recessive disorder characterized by cerebellar ataxia, ID, and disequilibrium syndrome (CAMRQ, MIM 615268), or severe hypotonia, ID, and optic atrophy with or without encephalopathy ^32–36^. A *de novo* balanced translocation leading to haploinsufficiency of this gene has been also proposed as the cause of moderate ID and hypotonia ^37^.

Downregulation of *ATP9A* has been associated with a significant increase of extracellular vesicles release, in particular the exosome ^15,16^. Extracellular vesicles release is an important form of intercellular communication that enables the transport of several different signaling molecules - including proteins and RNA – without the need of direct cell-to-cell contacts. It is involved in a wide range of biological processes, such as blood coagulation and immune response ^38,39^. Different physiological roles in the central nervous system have been proposed for extracellular vesicles, including neurite outgrowth and neuronal survival ^38,40^. Synaptic glutaminergic activity regulates the exosome release, pinpointing a role in maintenance of synaptic physiology ^41^. Interestingly, an increase of neuronal-derived exosome has been documented in individuals with Down syndrome as a compensatory mechanism for alterations in the endosomal pathway ^42,43^. Alteration in the recycling endosomal processes have been associated with Christianson syndrome (MIM 300243), a neurodevelopmental disorder characterized by ID, speech impairment, epilepsy, postnatal microcephaly, truncal ataxia, and hyperactivity ^44,45^. This syndrome is caused by pathogenic variants in the endosomal sodium hydrogen exchanger *SLC9A6* ^44,46–48^. Suggestive of neuronal degeneration, primary hippocampal neurons of *Slc9a6-/-* mouse or expressing a deleterious mutation in this gene presented with an increase of caspase activation and a decrease in dendrites’ lengths, size, and in dendritic arborization ^44,46,49^. ATP9A is known to be involved in the endosomal recycling pathway ^12,15,16^. For instance, depletion of ATP9A reduces the plasma membrane expression of the glucose transporter GLUT1 and increases its level in endosome, altering its recycling ^12^. Deficiency of GLUT1 has been associated with a neurological disorder with a variable phenotype including epilepsy, movement disorders, mild to severe intellectual disability, and acquired microcephaly in some cases ^50,51^. Furthermore, a gene expression microarray analysis performed on knockdown *ATP9A* HepG2 cells has shown that the differentially expressed genes are involved in endocytosis ^15^.

Using the data of the Bravo database, we have attempted to estimate the number of individuals affected by *ATP9A*-related neurodevelopmental disorder. There are 624 deleterious alleles in Bravo (Loss of function and non-synonymous variants with CADD>25). Thus, the allelic frequency of likely pathogenic *ATP9A* variants is estimated to be 1 in 424 and the frequency of heterozygous carriers 1 in 212. Using these frequencies, we estimated births per year in outbred population and consanguineous marriages to be approximately 722 in 130,000,000 and 958 in 6,500,000 births, respectively. The total estimated number of new patients per year if 1680.

In conclusion, we describe a novel autosomal recessive neurodevelopmental disorder. In two unrelated consanguineous families, we identified variants predicted to affect the splicing of *ATP9A*. The three individuals homozygous for these putatively truncating variants presented with severe ID, motor and speech impairment, and behavioral anomalies. Consistent with a causative role of ATP9A in the patients’ phenotypes *Atp9a-/-*mouse model showed behavioral changes.

## METHODS

The current study was approved by the IRBs of the Khyber Medical University, Peshawar, Pakistan, and the University Hospitals of Geneva, Switzerland (Protocol number: CER 11-036). Informed consent forms were obtained from guardians of all affected individuals who participated in this study.

The proband IV:1 of family 1 was subjected to exome sequencing (ES). DNA was enriched using SureSelect Human All Exon v6 capture kit (Agilent Technologies, Santa Clara, CA, USA) and sequenced on an Illumina HiSeq 4000 platform, with an average coverage of 120x at each nucleotide position. ES data were analyzed with an in-house customized pipeline^8^ that is based on published algorithms including BWA, SAMtools ^52^, PICARD (http://broadinstitute.github.io/picard/) and (GATK) ^53^. Initial screening for known or novel pathogenic mutations in the reported ID genes was performed. The 720K SNP array was performed in parents (III:3 and III:4), affected (IV:1 and IV:7) and unaffected individuals (IV:3 and IV:5) of family 1 to identify Runs of Homozygosity (ROH) using PLINK as described previously ^54–56^. ROH and exome sequencing data were analyzed with CATCH ^57^ to determine variants that were present in ROHs of patients (IV:1 and IV:7) but not in normal individuals of family 1. Subsequently the variants were filtered manually by using the criteria described in published studies ^55,56^.

The exome of IV:1 from family 2 was captured using the xGen Exome Research Panel v2 (Integrated DNA Technologies) and sequenced using the Illumina HiSeq4000 platform according to the manufacturer’s protocols. The overall mean-depth base coverage was 153-fold and 97% of the targeted region was covered at least 20-fold. Read mapping and variant calling were performed as described ^58^ using the Varapp software ^59^. Homozygous and hemizygous variants with a MAF < 1% in the general population (1000genome, EVS, gnomAD) were retained and screened for variants in reported ID genes (Supplementary Table 1). Homozygosity mapping was performed with AutoMap, which uses Variant Call Format (VCF) files from WES ^60^.

## Data Availability

The data that support the findings of this study are available from the corresponding author upon request.

## DATA AVAILABILITY

The data that support the findings of this study are available from the corresponding authors upon request.

## ACKNOWLEDGMENTS

We thank the probands and their families for their participation in this study. This work was supported by grants from the Swiss National Science Foundation (31003A_182632) and the Lejeune Foundation (JLF #1838) to AR and the Childcare Foundation to SEA.

## AUTHORS CONTRIBUTION

FM and AR wrote the manuscript with the help of SEA and MA. FM and MA performed WES, in silico analysis of the variant and Sanger sequencing. SAP, HMAB provided clinical information for family 1. HD, AT, SGF and MC collected genomic DNAs and clinical information for family 2. SEA, MA and AR supervised the study.

---

**Supplementary Table 1**: Homozygous variants identified by WES in proband IV:1 of family 2

